# Derivation and external validation of clinical prediction rules identifying children at risk of linear growth faltering (stunting) presenting for diarrheal care

**DOI:** 10.1101/2022.03.08.22271796

**Authors:** Sharia M. Ahmed, Ben J. Brintz, Patricia B Pavlinac, Lubaba Shahrin, Sayeeda Huq, Adam C. Levine, Eric J. Nelson, James A Platts-Mills, Karen L Kotloff, Daniel T Leung

**Affiliations:** Division of Infectious Diseases, University of Utah School of Medicine, Salt Lake City, UT, USA; Division of Epidemiology, University of Utah School of Medicine, Salt Lake City, UT, USA; Department of Global Health, Global Center for Integrated Health of Women, Adolescents and Children (Global WACh), University of Washington, Seattle, WA, USA; International Centre for Diarrhoeal Disease Research, Bangladesh, Dhaka, Bangladesh; Department of Emergency Medicine, Warren Alpert Medical School of Brown University, Providence, RI, USA; Department of Pediatrics and Environmental and Global Health, Emerging Pathogens Institute, University of Florida, Gainesville, FL, USA; Division of Infectious Diseases and International Health, University of Virginia, Charlottesville, VA, USA; Department of Pediatrics, Center for Vaccine Development, University of Maryland School of Medicine, Baltimore, MD, USA

## Abstract

**Background:** Nearly 150 million children under-5 years of age were stunted in 2020. We aimed to develop a clinical prediction rule (CPR) to identify children likely to experience additional stunting following acute diarrhea, to enable targeted approaches to prevent this irreversible outcome.

**Methodology:** We used clinical and demographic data from the Global Enteric Multicenter Study (GEMS) study to build predictive models of linear growth faltering (decrease of ≥0.5 or ≥1.0 in height-for-age z-score [HAZ] at 60 day follow-up) in children ≤59 months presenting with moderate-to-severe diarrhea (MSD), and community controls, in Africa and Asia. We screened variables using random forests, and assessed predictive performance with random forest regression and logistic regression using 5-fold cross-validation. We used the Etiology, Risk Factors, and Interactions of Enteric Infections and Malnutrition and the Consequences for Child Health and Development (MAL-ED) study to A) re-derive, and B) externally validate our GEMS-derived CPR.

**Results:** Of 7639 children in GEMS, 1744 (22.8%) experienced severe growth faltering (≥0.5 decrease in HAZ). In MAL-ED, we analyzed 5683 diarrhea episodes from 1322 children, of which 961(16.9%) episodes experienced severe growth faltering. Top predictors of growth faltering in GEMS were: age, HAZ at enrollment, respiratory rate, temperature, and number of people living in the household. The maximum AUC was 0.75 (95% CI: 0.75, 0.75) with 20 predictors, while 2 predictors yielded an AUC of 0.71 (95% CI: 0.71, 0.72). Results were similar in the MAL-ED re-derivation. A 2-variable CPR derived from children 0-23 months in GEMS had an AUC=0.63 (95% CI 0.62, 0.65), and AUC=0.68 (95% CI: 0.63, 0.74) when externally validated in MAL-ED.

**Conclusions:** Our findings indicate that use of prediction rules could help identify children at risk of poor outcomes after an episode of diarrheal illness.

## INTRODUCTION

Despite recent advances in the prevention and treatment of childhood malnutrition, nearly 150 million children under-5 years of age were stunted in 2020(1). Stunting is defined as a length- or height-for-age z-score 2 or more standard deviations below the population median(2), and is considered both an indicator of underlying deficits (i.e. chronic malnutrition(3)), as well as a potential contributor to future health problems (e.g. through poor immune system maturation(4, 5)). Furthermore, stunting has been consistently associated with increased risk of morbidity and mortality, delayed or deficient cognitive development, and reduced educational attainment(6-12). Timely and accurate identification of children most likely to experience stunting offers an opportunity to prevent such negative health outcomes.

Stunting has been linked with diarrheal diseases across many settings(13). An estimated 10.9% of global stunting is attributable to diarrhea(14), and a child with diarrhea is more likely to have a lower HAZ score or to die than age-matched controls(15). Given the 1.1 billion episodes of childhood diarrhea that occur globally every year(16), assessment of children seeking healthcare for diarrhea treatment provides an opportunity to identify those at increased risk for negative outcomes, including stunting and death. Once identified, these children could be specifically targeted for intensive interventions, thereby more efficiently allocating public health resources.

In this study, we aimed to develop parsimonious, easy to implement clinical prediction rules (CPRs) to identify children under-5 most likely to experience linear growth faltering among community-dwelling children presenting to care for acute diarrhea. CPRs are algorithms that aid clinicians in interpreting clinical findings and making clinical decisions(17). Linear growth faltering, or falling below standardized height/length growth trajectory projections, captures children whose growth has slowed precipitously and is a precursor of stunting. A number of prior studies have identified risk factors for linear growth faltering(14, 18-26), but many of these were single-site studies using traditional model building approaches, some of which lacked appropriate assessments of model discrimination and calibration. Building on this body of literature, we used machine learning methods on data from two large multi-center studies to derive and externally validate prediction models for growth faltering, with the hopes of reliably identifying children that would most benefit from additional nutritional intervention after care for acute diarrhea.

## METHODS

### Study Population for Derivation Cohort 1 (GEMS)

We used data from The Global Enteric Multicenter Study (GEMS) to derive CPRs for growth faltering. The GEMS study has been described elsewhere in-depth(15, 27). Briefly, GEMS was a prospective case-control study of acute moderate to severe diarrhea (MSD) in children 0-59 months of age. Data were collected in December 2007 – March 2011 from 7 sites in Africa and Asia, including those in Mali, The Gambia, Kenya, Mozambique, Bangladesh, India, and Pakistan. MSD was defined as diarrhea accompanied by one or more of the following: dysentery, dehydration, or hospital admission. Diarrhea was defined as new onset (after ≥7 days diarrhea-free) of 3 or more looser than normal stools in the previous 24 hours lasting 7 days or less. Cases were enrolled at initial presentation to a sentinel hospital or health center, and matched within 14 days to 1-3 controls without diarrhea enrolled from the community. Demographics, epidemiological, and clinical information was collected from caregivers of both cases and controls via standardized questionnaires, and clinic staff conducted physical exams and collected stool samples which have undergone conventional and molecular testing to ascertain the pathogen that caused the diarrhea. Approximately 60 days (up to 91) after enrollment, fieldworkers visited the homes of both cases and controls to collect standardized clinical and epidemiological information and repeat anthropometry.

Children were excluded if follow-up observations occurred <49 or >91 days after enrollment, or if HAZ measurements were implausible (28), defined as: a) HAZ>6 or HAZ<-6; b) change in HAZ>3; c) >1.5cm loss of height from enrollment to follow-up; d) growth of >8cm or >4cm at 49-60 day follow-up for children ≤6 months and >6 months old, respectively; e) growth >10cm or >6cm at 61-91 day follow-up for children ≤6 months and >6 months old, respectively.

Parents or caregivers of participants provided informed consent, either in writing or witnessed if parents or caregivers were illiterate. The GEMS study protocol was approved by ethical review boards at each field site and the University of Maryland, Baltimore, USA.

### Study Population for Derivation Cohort 2 (MAL-ED)

We used the Etiology, Risk Factors, and Interactions of Enteric Infections and Malnutrition and the Consequences for Child Health and Development (MAL-ED) study to A) re-derive the best full model, and B) externally validate a 2-variable parsimonious version of our GEMS-derived CPR for growth faltering. MAL-ED is a longitudinal birth cohort study, and study details have been described elsewhere (29-32). In brief, healthy children were enrolled within 17 days of birth and followed prospectively through 24 months of age. Children were enrolled from October 2009 – March 2012 from 8 countries in Asia, Africa, and South America, including Tanzania, South Africa, Pakistan, India, Nepal, Bangladesh, Peru, and Brazil. Information on household, demographic, and clinical data from mother and child were collected at enrollment and reassessed periodically, and illness and feeding information was collected at twice-weekly household visits.

In MAL-ED, diarrhea was defined as maternal report of three or more loose stools in a 24 hour period, or one loose stool with blood. Each diarrhea episode had to be separated by at least 2 days without diarrhea in order to qualify as distinct diarrhea episodes. To match MAL-ED longitudinal cohort active surveillance data to GEMS, in which children were enrolled upon presentation to clinic with acute diarrhea, we linked anthropometric measurements and other predictor variables with diarrhea episodes in MAL-ED using the following methods (https://github.com/LeungLab/CPRgrowthfaltering): First, each episode of diarrhea was linked to the closest HAZ measurement from before the onset of diarrhea symptoms, but no more than 31 days beforehand. Each diarrhea episode was also linked with the HAZ measurement closest to 75 days after the onset of diarrhea symptoms, but within 49 and 91 days inclusive. Second, each diarrhea episode was linked to the closest observation of each potential predictor variable. Each dietary intake variable had to be observed within 90 days of the diarrhea episode, and each household descriptor variable had to be observed within 6 months of the onset of diarrhea in order to be eligible, otherwise those predictors were considered missing for that specific diarrhea episode. Finally, data were split into age categories, and only one diarrhea episode per enrolled child per model was randomly selected without replacement for analysis.

The same inclusion/exclusion criteria were applied as listed above for the GEMS growth faltering analysis, with the exception that the allowed follow-up period extended up to and including 95 days.

Parents or caregivers of participants provided informed consent. The MAL-ED study protocol was approved by ethical review boards at each field site and the Johns Hopkins Institutional Review Board, Baltimore, USA.

### Outcomes

We defined growth faltering as a decrease in height-for-age z-score (HAZ) of ≥0.5 HAZ within 49-91 days of enrollment in GEMS, or within 49-95 days in MAL-ED.

### Predictive Variables

In GEMS, potential predictors included over 130 descriptors of the child, household, and community, collected at enrollment (Supplemental Table S1). Collinear or conceptually similar predictors were removed from consideration to maximize model utility (e.g. HAZ, but not MUAC was considered in the main model). We considered individual components of household wealth, but did not explore the composite wealth variable used in other reports (28) since its utilization in a CPR would require collecting multiple parameters that were already being considered individually.

In MAL-ED, we considered 60 potential predictors of growth faltering (Supplemental Table S1). We limited possible predictor variables to those that would be easily assessable upon presentation to clinic in a low-resource setting (i.e. did not consider characteristics that required diagnostic testing), and again only considered individual components of combination indicators (e.g. wealth index, Vesikari score).

### Statistical Analysis

We screened variables using variable importance measures from random forests to identify the most predictive variables. Random forests are an ensemble learning method whereby multiple decision trees (1000 throughout this analysis) are built on bootstrapped samples of the data with only a random sample of potential predictors considered at each split, thereby decorrelating the trees and reducing variability(33). Throughout this analysis, the number of variables considered at each split was equal to the square root of the total number of potential variables, rounded down. Variables were ranked by predictive importance based on the reduction in mean squared prediction error achieved by including the variable in the predictive model on out-of-bag samples (i.e. observations not in the bootstrapped sample).

Generalizable performance was assessed using 5-fold repeated cross-validation. In each of 100 iterations, random forests were fit to a training dataset (random 80% sample of analytic dataset), and variable were ranked using the random forest importance measure as above. Separate logistic regression and random forest regression models were then fit to a subset of the top predictive variables in the training dataset. Subsets examined were the top 1-10, 15, 20, 30, 40, and 50 predictors. Each of these models were then used to predict the outcome (growth faltering) on the test dataset. Model performance was assessed using the receiver operating characteristic (ROC) curves and the cross-validated C-statistic (area under the ROC curve (AUC)), a measures which describes how well a model can discriminate between the two outcomes, from the cross-validation.

We assessed model calibration both quantitatively and graphically (“weak” and “moderate” calibration, respectively(34)). First, we assessed calibration-in-the-large, or calibration intercept, by using logistic regression to estimate the mean while subtracting out the estimate (model the log-odds of the true status, offset by the CPR-predicted log-odds). Next, we used calibration slope to assess the spread of the estimated probabilities, whereby we fit a logistic regression model with log-odds of the true status as the dependent variable and CPR-predicted log-odds as the independent variable. Finally, we assessed moderate calibration graphically, whereby we calculated the predicted probability of growth faltering for each child in a given analysis using each iteration of each n-variable model fit. These predicted probabilities were then binned into deciles, and the proportion of each decile who truly experienced the outcome was calculated for each iteration of each n-variable model. The mean predicted probability and observed proportion was calculated for each decile across iterations. These average observed proportions were then plotted against averaged deciles for each n-variable model fit (see https://github.com/LeungLab/CPRgrowthfaltering for full analytic code).

Based on top predictors available in both GEMS and MAL-ED (see Results), the 2-variable GEMS-derived CPR of growth faltering was externally validated in MAL-ED data. A logistic regression was fit to all diarrhea cases age 0-23 months in GEMS data, with predictors chosen based on random forest. This model was then used to predict growth faltering in diarrhea cases in MAL-ED (age in MAL-ED converted from days to months), and discrimination and calibration were assessed as described above.

### Sensitivity and Subgroup Analyses

We undertook additional sensitivity and subgroup analyses to explore if our ability to predict growth faltering improved in specific patient populations or with additional predictors within GEMS data. First, we explored age-strata specific CPRs for children 0-11months, 12-23months, 0-23months, and 24-59 months. Second, we explored the predictive ability of MUAC instead of and in addition to HAZ. Third, we attempted to account for potential seasonal variation by adding a predictor for month of diarrhea. Fourth, we added indicator variables for the use of antibiotics before presentation (enrollment), while at clinic, prescription to take home after care, and ever. Fifth, we limited our outcome to only very severe growth faltering, defined as a decrease ≥1.0 HAZ (as opposed to ≥0.5 HAZ in the main analysis). Sixth, we explored the impact diarrhea etiology had on growth faltering prediction. We added variables for the presence/absence of *Shigella, Cryptosporidium, Shigella* + *Cryptosporidium* infections, and any viral etiology (defined as infection of any of the following: astrovirus, norovirus GII, rotavirus, sapovirus, and adenovirus 40/41). Etiology-specific infection were defined as an episode-specific attributable fraction (AFe) greater than or equal to a given cutoff (0.3, 0.5, and 0.7 were considered)(15). Finally, we explored the prevalence of growth faltering in healthy controls, and identified top predictors and their ability to predict growth faltering in controls. Potential predictors related to diarrhea were not considered amongst controls (e.g. number of days with diarrhea at presentation).

## RESULTS

### Growth faltering in children following acute diarrhea in GEMS and MAL-ED

There were 9439 children with acute diarrhea enrolled in GEMS. In the analysis of the primary outcome (growth faltering), 110 observations were dropped for having follow-up measurements taken <49 or >91 days after enrollment, and 1276 were dropped for having implausible or missing HAZ measurements, leaving an analytic sample of 8053. An addition 414 observations were dropped for having missing predictor data, as random forest analysis requires complete cases. Of the remaining 7639 children, 1744 (22.8%) experienced severe growth faltering (≥0.5 decrease in HAZ), and 357 (4.7%) experienced very severe growth faltering (≥1.0 decrease in HAZ) (Supplemental Figures S1). Growth faltering rates differed by country, with Mozambique and The Gambia having the highest rates of growth faltering (34.5% and 31.9% experienced severe growth faltering, respectively) and Mali having the lowest rate (14.9%, Supplemental Table S2). Growth faltering rates also varied by child’s age, with a higher proportion of younger children experiencing growth faltering than older children (Supplemental Table S3).

In the analysis of MAL-ED data, we started with 6617 diarrhea episodes from 1390 children. In order to align with GEMS inclusion criteria and limit to acute onset diarrhea, 566 diarrhea episodes were dropped for having prolonged or persistent diarrhea (>7 days duration). An additional 125 episodes were dropped for having missing HAZ measurements or an HAZ follow-up measurement <49 or >95 days from diarrhea onset, and 138 episodes were dropped for having implausible HAZ measurements, leaving 5788 diarrhea episodes from 1350 children. An additional 105 observations were dropped for having missing predictor data. Of the remaining 5683 observations from 1322 children, 961(16.9%) episodes experienced severe growth faltering (≥0.5 decrease in HAZ) and 161(2.8%) episodes experienced very severe growth faltering (≥1.0 decrease in HAZ, Figure S1).

### Derivation of a CPR to identify children who went on to severe growth faltering following acute diarrhea using GEMS data

After random forest screening of variables, logistic regression models consistently had higher AUCs than random forest regression models (Supplementary Figure S2), therefore we only present the easier to interpret logistic regression results moving forward. In Table 1, we show the top-10 most predictive variables ranked from most to least important, for severe growth faltering (≥0.5 decrease in HAZ) and death, respectively. The top predictive variables for severe growth faltering were: age, HAZ at enrollment, respiratory rate, temperature, number of people living in the household, number of people sleeping in the household, number of days of diarrhea at presentation, number of other households that share same fecal waste disposal facility (e.g. latrine), whether the child was currently breastfed at time of diarrhea, and the number of children <60 months old living in the household. The maximum AUC attained with the model was 0.75 (94% CI: 0.75, 0.75) with a model of 20 variables, while an AUC of 0.71 (95% CI: 0.71, 0.72), 0.72 (95% CI: 0.72, 0.72), and 0.72 (95% CI: 0.72, 0.72) could be obtained with a CPR of 2, 5, and 10 variables, respectively (Supplemental Figure S2). When limited to children 0-23 months of age, AUC decreased to 0.64 (95% CI: 0.64, 0.64) for 10 variables. In the full 10-variable model, we achieved a specificity of 0.47 at a sensitivity of 0.80 (Figure 1). The average predicted probability of growth faltering was consistently close to the average observed probability (calibration-in-the-large, or intercept), and the spread of predicted probabilities was similar to the spread of observed probabilities (calibration slope) for models including 1 to 10 predictor variables (Table 2, Figure 2).

**Table 1:** GROWTH FALTERING: Variable importance ordering and cross-validated average overall AUC and AUC by patient subset and 95% confidence intervals for a 5 (bold) and 10 (italicized) variable logistic regression model for predicting growth faltering in children in 7 LMICs derived from GEMS data (≥0.5 decrease in HAZ in children with acute diarrhea)

|  | GEMS |  |  | MAL-ED |
| --- | --- | --- | --- | --- |
| Variable/<br>Patient<br>Subset | 0-59mo<br>(main text<br>model) | 0-23mo (for<br>external<br>validation) | Healthy<br>controls | 0-23mo |
| AUCs | **0.72 (0.72,**<br>**0.72)** | **0.64 (0.63,**<br>**0.65)** | **0.79 (0.78,**<br>**0.79)** | **0.67 (0.67,**<br>**0.68)** |
|  | *0.72 (0.72,*<br>*0.72)* | *0.64 (0.64,*<br>*0.64)* | *0.79 (0.79,*<br>*0.79)* | *0.68 (0.67,*<br>*0.69)* |
| 1 | Age<br>(months) | HAZ | Age<br>(months) | HAZ |
| 2 | HAZ | Age<br>(months) | HAZ | Age (days) |
| 3 | Respiratory<br>rate | Temperature | Respiratory<br>rate | Total days<br>breastfeeding |
| 4 | Temperature | Respiratory<br>rate | Temp | Total days in<br>all diarrheal<br>episodes to<br>date |
| 5 | Num.<br>people<br>living in<br>household | Num.<br>people<br>living in<br>household | Num.<br>people<br>living in<br>household | Mean<br>number of<br>people per<br>room |
| 6 | Num. rooms<br>used for<br>sleeping | Num. rooms<br>used for<br>sleeping | Breastfed | Days with<br>diarrhea so<br>far in this<br>episode |
| 7 | Num. days<br>of diarrhea<br>at<br>presentation | Num. days<br>of diarrhea<br>at<br>presentation | Num.<br>rooms used<br>for<br>sleeping | Maternal<br>education<br>(years) |
| 8 | Num. other households that share same fecal waste facility | Num. other households that share same fecal waste facility | Num. children <60months live in household | Days since last diarrhea episode |
| 9 | Breastfed | Num. children <60 months live in household | Caregiver education | People sleeping in house |
| 10 | Num. children <60months live in household | Caregiver education | Num. other households share latrine | Max loose stools in this episode |

**Figure 1:**
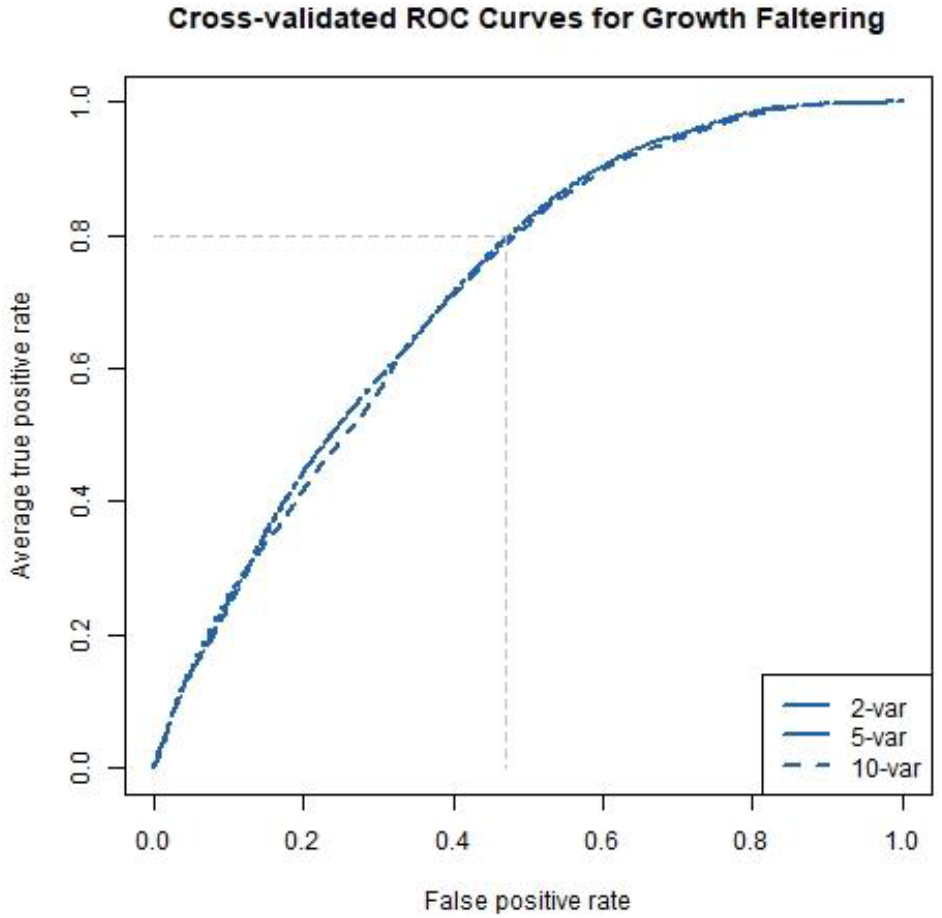
ROC curves: Average ROC curves from the cross-validated logistic regression models predicting growth faltering with 2, 5, and 10 predictors. The faded dashed lines represent specificity (1-false positive rate) achievable with a sensitivity (true positive rate) of 0.80 for prediction of the outcome.

**Table 2:**
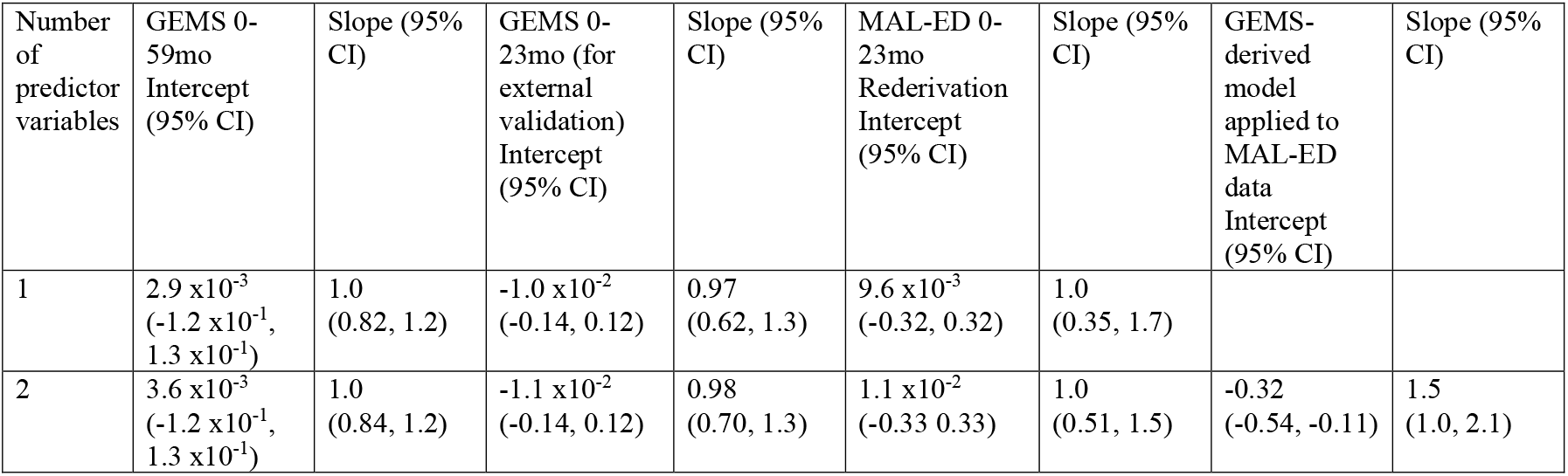
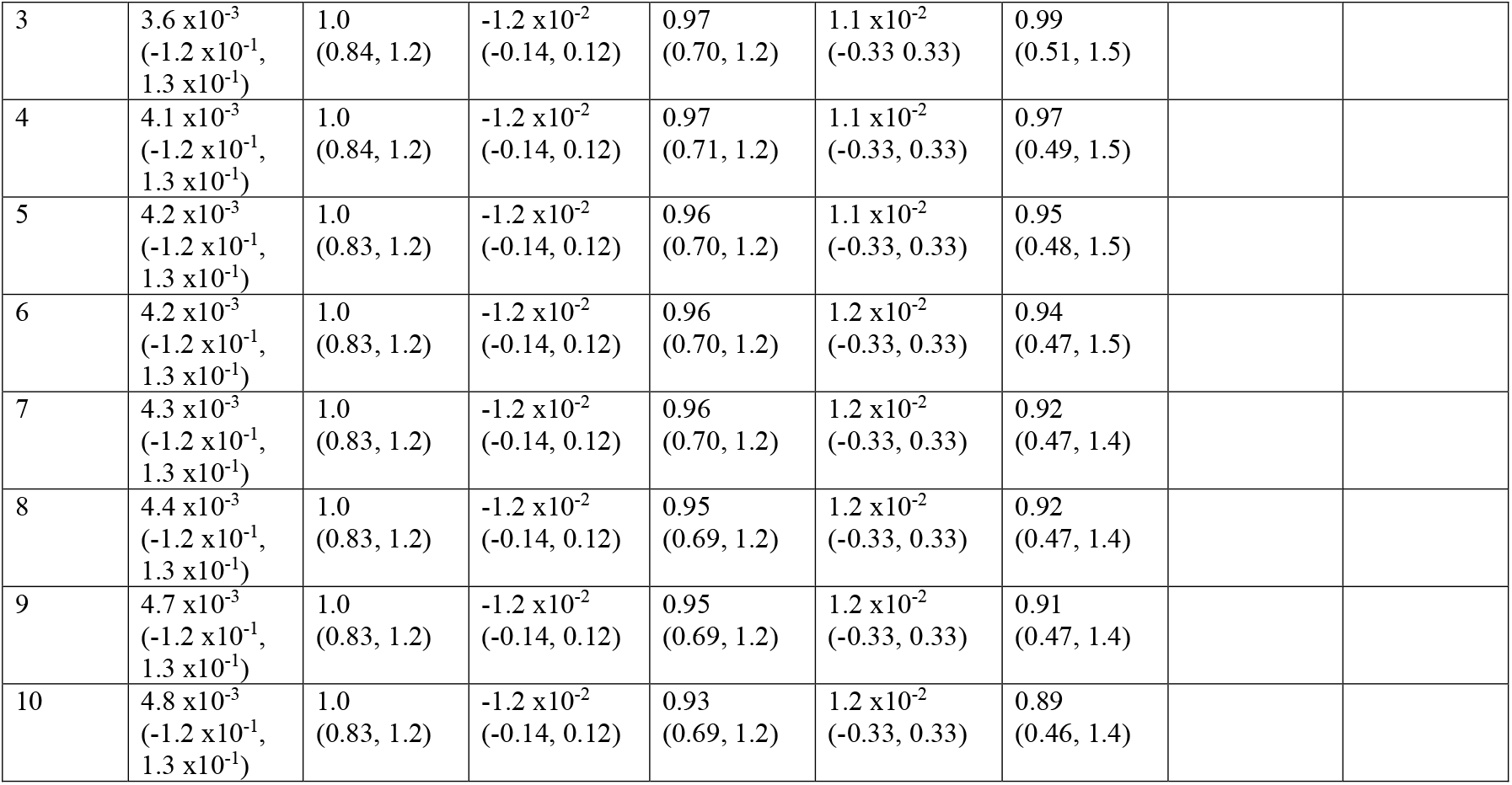
Calibration intercept and slope

| Number of predictor variables | GEMS 0-59mo Intercept (95% CI) | Slope (95% CI) | GEMS 0-23mo (for external validation) Intercept (95% CI) | Slope (95% CI) | MAL-ED 0-23mo Rederivation Intercept (95% CI) | Slope (95% CI) | GEMS-derived model applied to MAL-ED data Intercept (95% CI) | Slope (95% CI) |
| --- | --- | --- | --- | --- | --- | --- | --- | --- |
| 1 | $2.9 \times 10^{-3}$<br>($-1.2 \times 10^{-1}$, $1.3 \times 10^{-1}$) | 1.0<br>(0.82, 1.2) | $-1.0 \times 10^{-2}$<br>(-0.14, 0.12) | 0.97<br>(0.62, 1.3) | $9.6 \times 10^{-3}$<br>(-0.32, 0.32) | 1.0<br>(0.35, 1.7) | | |
| 2 | $3.6 \times 10^{-3}$<br>($-1.2 \times 10^{-1}$, $1.3 \times 10^{-1}$) | 1.0<br>(0.84, 1.2) | $-1.1 \times 10^{-2}$<br>(-0.14, 0.12) | 0.98<br>(0.70, 1.3) | $1.1 \times 10^{-2}$<br>(-0.33 0.33) | 1.0<br>(0.51, 1.5) | -0.32<br>(-0.54, -0.11) | 1.5<br>(1.0, 2.1) |
| 3 | $3.6 \times 10^{-3}$<br>($-1.2 \times 10^{-1}$,<br>$1.3 \times 10^{-1}$) | 1.0<br>(0.84, 1.2) | $-1.2 \times 10^{-2}$<br>(-0.14, 0.12) | 0.97<br>(0.70, 1.2) | $1.1 \times 10^{-2}$<br>(-0.33, 0.33) | 0.99<br>(0.51, 1.5) | | |
| 4 | $4.1 \times 10^{-3}$<br>($-1.2 \times 10^{-1}$,<br>$1.3 \times 10^{-1}$) | 1.0<br>(0.84, 1.2) | $-1.2 \times 10^{-2}$<br>(-0.14, 0.12) | 0.97<br>(0.71, 1.2) | $1.1 \times 10^{-2}$<br>(-0.33, 0.33) | 0.97<br>(0.49, 1.5) | | |
| 5 | $4.2 \times 10^{-3}$<br>($-1.2 \times 10^{-1}$,<br>$1.3 \times 10^{-1}$) | 1.0<br>(0.83, 1.2) | $-1.2 \times 10^{-2}$<br>(-0.14, 0.12) | 0.96<br>(0.70, 1.2) | $1.1 \times 10^{-2}$<br>(-0.33, 0.33) | 0.95<br>(0.48, 1.5) | | |
| 6 | $4.2 \times 10^{-3}$<br>($-1.2 \times 10^{-1}$,<br>$1.3 \times 10^{-1}$) | 1.0<br>(0.83, 1.2) | $-1.2 \times 10^{-2}$<br>(-0.14, 0.12) | 0.96<br>(0.70, 1.2) | $1.2 \times 10^{-2}$<br>(-0.33, 0.33) | 0.94<br>(0.47, 1.5) | | |
| 7 | $4.3 \times 10^{-3}$<br>($-1.2 \times 10^{-1}$,<br>$1.3 \times 10^{-1}$) | 1.0<br>(0.83, 1.2) | $-1.2 \times 10^{-2}$<br>(-0.14, 0.12) | 0.96<br>(0.70, 1.2) | $1.2 \times 10^{-2}$<br>(-0.33, 0.33) | 0.92<br>(0.47, 1.4) | | |
| 8 | $4.4 \times 10^{-3}$<br>($-1.2 \times 10^{-1}$,<br>$1.3 \times 10^{-1}$) | 1.0<br>(0.83, 1.2) | $-1.2 \times 10^{-2}$<br>(-0.14, 0.12) | 0.95<br>(0.69, 1.2) | $1.2 \times 10^{-2}$<br>(-0.33, 0.33) | 0.92<br>(0.47, 1.4) | | |
| 9 | $4.7 \times 10^{-3}$<br>($-1.2 \times 10^{-1}$,<br>$1.3 \times 10^{-1}$) | 1.0<br>(0.83, 1.2) | $-1.2 \times 10^{-2}$<br>(-0.14, 0.12) | 0.95<br>(0.69, 1.2) | $1.2 \times 10^{-2}$<br>(-0.33, 0.33) | 0.91<br>(0.47, 1.4) | | |
| 10 | $4.8 \times 10^{-3}$<br>($-1.2 \times 10^{-1}$,<br>$1.3 \times 10^{-1}$) | 1.0<br>(0.83, 1.2) | $-1.2 \times 10^{-2}$<br>(-0.14, 0.12) | 0.93<br>(0.69, 1.2) | $1.2 \times 10^{-2}$<br>(-0.33, 0.33) | 0.89<br>(0.46, 1.4) | | |

**Figure 2:**
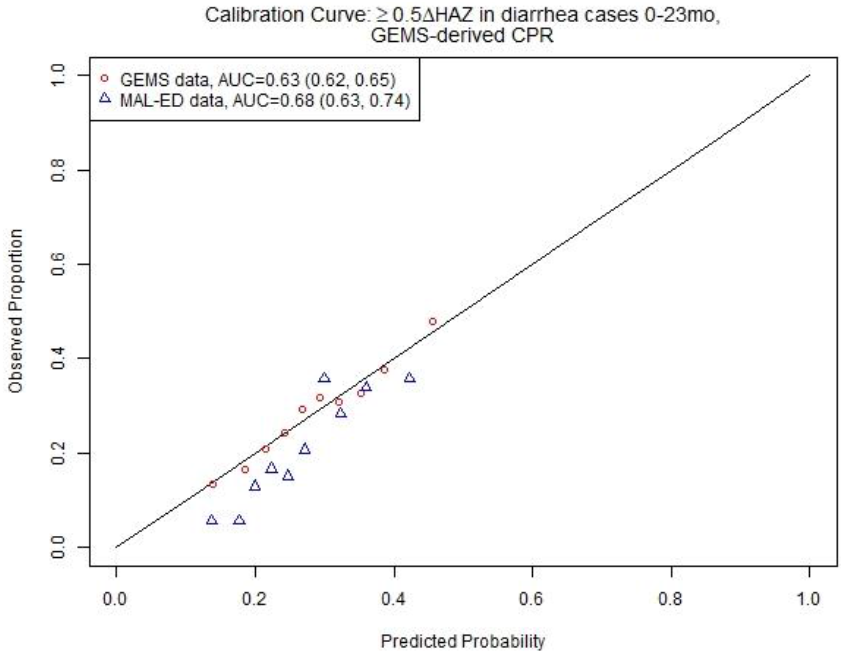
2-Variable CPR for growth faltering: Calibration curve and discriminative ability of 2-varaible (age, HAZ at enrollment) model predicting growth faltering (≥0.5 decrease in HAZ) in children presenting for acute diarrhea in LMICs.

### Rederivation and external validation of a CPR to identify children who went on to severe growth faltering following acute diarrhea using MAL-ED data

We then derived a CPR for growth faltering using MAL-ED data, and found that the top predictors were similar to those identified using GEMS data, with age and HAZ at diarrhea being the top two predictors. Other top predictors in MAL-ED included breastfeeding, total days in all diarrhea episodes, mean number of people per room of home, days with diarrhea so far in this episode, number of years of maternal education, days since last diarrhea episode, number of people sleeping in house, and loose stools in this diarrhea episode (Table 1). The discriminative performance of the full model was similar to that found with GEMS (0.72 (95% CI: 0.72, 0.72) in GEMS, 0.68 (95% CI: 0.67, 0.69) in MAL-ED). The average predicted probability of growth faltering was consistently close to the average observed probability (calibration-in-the-large, or intercept). The spread of predicted probabilities (calibration slope) was slightly more extreme than observed probabilities, but there was no evidence they were different than 1.0 for models including 1 to 10 predictor variables (slope point estimates all 95% CI include 1.0, see Table 2, Supplemental Figure S3).

Due to a lack of overlap in variables between datasets, we were unable to externally validate the 10-variable version of our growth faltering CPR. However, the top two predictors were available in both the GEMS and MAL-ED dataset. Therefore, we took the 2-variable CPR of growth faltering derived from children 0-23 months of age in GEMS, including HAZ at enrollment and age (the top two predictors), and externally validated it in MAL-ED data. The CPR had marginal discrimination in the GEMS data (AUC=0.64, 95% CI 0.64, 0.64), and a slight increase in discriminative ability at external validation in MAL-ED data (AUC=0.68, 95% CI: 0.63, 0.74). On average, the CPR overestimated probability of growth faltering (calibration intercept −0.32, 95% CI: − 0.54, −0.11), and predictions tended to be too moderate (calibration slope 1.5, 95% CI: 1.0, 2.1) (Table 2, Figure 2). Odds ratios for the 10-variable model predicting severe growth faltering in MAL-ED are shown in Supplemental Table S4.

### Addition of MUAC, diarrhea etiology, and antibiotic use did not meaningfully impact discriminative performance of CPR to identify children who went on to severe growth faltering following acute diarrhea in GEMS

Table 1 and Supplemental Table S5 present the results of the growth faltering sensitivity analyses. Top predictor variables were highly consistent across models and included patient demographics, patient symptoms, and indicators of household wealth. CPR’s of higher age strata had higher AUCs (0.76 (95% CI: 0.75, 0.77) in 24-59mo in GEMS versus 0.60 (95% CI: 0.59, 0.60) in 0-11mo in GEMS).

When MUAC was considered as a potential predictor (instead of HAZ), MUAC replaced HAZ as a top predictor, all other top-10 predictors remained the same, and AUC decreased (down to 0.70, 95% CI: 0.70, 0.70). When both HAZ and MUAC were considered as potential predictors, both were top predictors, but AUC remained unchanged compared to the main model that considered only HAZ (0.72, 95% CI: 0.72, 0.73). The predictors of very severe growth faltering (≥1.0 decrease in HAZ) were similar to the predictors of severe growth faltering (≥0.5 decrease in HAZ), though predictive ability was better (AUC 0.80 (95% CI: 0.79, 0.80) for ≥1.0 versus 0.72 (95% CI: 0.71, 0.73) for ≥0.5).

Accounting for seasonality did not meaningfully improve the CPR, and antibiotic use and diarrhea etiology were consistently not ranked as top predictors of growth faltering (Supplemental Table S5). Finally, including more predictor variables only marginally improved AUCs.

### Derivation of a CPR to identify children without diarrhea (controls) who went on to severe growth faltering using GEMS data

Top predictors of growth faltering were similar in non-diarrhea controls compared to cases in GEMS (Table 1), but predictive ability was higher (AUC 0.79 (95% CI: 0.78, 0.79) in controls versus 0.72 (95% CI: 0.72, 0.72) in cases). Again, top predictors were consistent with previous models and included age, HAZ at enrollment, respiratory rate, temperature, number of people living in household, breastfed, number of rooms used for sleeping, number of children under 60 months old who live in household, education level of primary caregiver, and number of other households that share same fecal waste disposal facility (e.g. latrine). The maximum AUC attained with the model was 0.79 (95% CI: 0.79, 0.80) with a model of 15 variables, while an AUC of 0.79 (95% CI: 0.78, 0.79) and 0.79 (95% CI: 0.79, 0.79) could be obtained with a CPR of 5 and 10 variables, respectively (Supplemental Figure S2).

## DISCUSSION

By utilizing data from two large multi-center clinical studies of pediatric diarrhea, we used a combination of machine learning and conventional regression methods to derive and validate clinical prediction rules for linear growth faltering. The discriminative performance of our CPR for growth faltering was remarkably similar between the two datasets (AUC=0.72, 95% CI: 0.72, 0.72, based on GEMS 0-59 months; 0.68, 95% CI: 0.67, 0.69 based on MAL-ED 0-24 months). We were then able to externally validate a 2-variable version, which also had similar discriminative ability between the datasets (AUC 0.64 to 0.68 for 0-23 and 0-24 months in GEMS and MAL-ED respectively). Our findings suggest the potential for a parsimonious prediction rule-guided algorithm to identify young children with acute diarrhea for appropriate triage and follow-up.

The limited number of studies that aim to identify children most likely to growth falter after acute diarrhea have resulted in CPRs with varying discriminative and generalizability. Our full CPRs were better at identifying growth faltering than Brander et al (28) (AUC=0.67, 95% CI: 0.64, 0.69), which was not externally validated, and worse than Hanieh(35) et al (AUC: 0.85, 95% CI 0.80, 0.90), which only used data from a single country. The top predictors of growth faltering identified by random forests in our analysis were consistent with existing knowledge of the drivers of growth faltering – child demographics, child symptoms, and indicators of household wealth. The top two variables (used in our parsimonious externally validated CPR) were age and baseline HAZ. However, despite the inclusion of markers of disease severity (temperature, respiratory rate, number of days of diarrhea), overall ability to predict growth faltering was moderate, and consideration of additional factors related to nature of disease (etiology, antibiotics) did not improve discriminative ability. This is consistent with previous analysis in GEMS data that found treating diarrhea with antibiotics generally did not prevent growth faltering (except for Shigella infections(36)).

Furthermore, the similar incidence of growth faltering in diarrhea cases and matched controls (particularly in the youngest children), as well as the almost identical predictive variables and similar AUCs, suggests that the impact of a single episode of acute diarrhea on growth trajectory may be relatively low. It is possible that the entire diarrheal history of a child (e.g. frequency and severity of acute diarrhea), or subclinical enteric infections that do not result in diarrhea, are more important to their growth trajectory than a single diarrheal episode, though evidence is mixed(13, 26, 37). Indeed, the average baseline HAZ at enrollment was 0.5 HAZ lower in children who did not experience growth faltering than in children who did (Supplemental Figure S4), suggesting the possibility that children need to have high enough HAZ in order to have the potential to falter. It is also possible that the underlying cause(s) of stunting are complex and interrelated, and relatively simple predictive models are not able to accurately parse apart which children do and do not experience sufficient causes.

While effective interventions exist for treating acute malnutrition (e.g. exclusive breastfeeding for the first 6 months of life, inpatient- and community-based management of acute malnutrition using corn-soy blend or ready-to-use therapeutic food (38-40)), there are few evidence-based guidance on how to reverse the effects of chronic malnutrition once a child is stunted(39, 41-44)). We found that approximately 1 in 5 children experience severe growth faltering subsequent to acute diarrhea, that is, an *additional* ≥0.5 decrease in HAZ in the 2-3 months after acute diarrhea. Currently, presenting to care for an acute illness, such as diarrhea, offers an opportunity for medical personnel to assess and treat children for acute malnutrition through intensive feeding programs. Our CPR provides a tool for identifying patients likely to experience additional growth faltering after acute diarrhea. This would allow clinicians to connect patients with community-based nutrition interventions (e.g. maternal support for safe introduction of weening foods, small quantity lipid nutrient supplements (SQ-LNS), etc.(45-48)) to prevent *additional* effects of chronic malnutrition, namely irreversible stunting.

Our study has a number of strengths and limitations. We derived CPRs for growth faltering from two multi-site, prospective studies that included longitudinal follow-up with extensive etiologic testing. Unlike previous work in this area, we used random forests for variable selection which do not require assumptions about the underlying variables and generally outperform(49) conventional model building techniques. We were able to re-derive the 10-variable version in two distinct datasets with similar results. While we were only able to externally validate a 2-variable version of our growth faltering CPR, its discriminative performance was similar to the full 10-variable version, and was robust to external validation. Furthermore, while the observation windows were large for many variables in the MAL-ED dataset used for external validation (up to 90 days for dietary variables, and up to 6months for household descriptors), the variables of interest in the 2-variable CPR were observed no more than 31 days from the start of diarrhea. In addition, we considered all diarrhea as an outcome of interest in MAL-ED, whereas the analysis in GEMS was limited to MSD. When limiting the MAL-ED analysis to MSD as defined in GEMS, the top predictors and discriminative ability were very similar. Finally, we explored a range of AFe cutoffs for etiology, with consistent results.

In conclusion, we used data from two large multi-country studies to derive and validate a clinical prediction rule for growth faltering in children presenting for diarrhea treatment. Our findings indicate that use of prediction rules, potentially applied as clinical decision support tools, could help to identify children at risk of poor outcomes after an episode of diarrheal illness. In settings with high mortality and morbidity in early childhood, such tools could represent a cost-effective way to target resources towards those who need it most.

## Data Availability

All data used in the present study are available from ClinEpiDB.org with approval of relevant study leads.

https://clinepidb.org/ce/app/record/dataset/DS_841a9f5259

https://clinepidb.org/ce/app/record/dataset/DS_5c41b87221

