## Supplemental File for "Derivation and external validation of clinical prediction rules identifying children at risk of linear growth faltering (stunting) presenting for diarrheal care"

Figure S1: Flow diagram of study inclusion

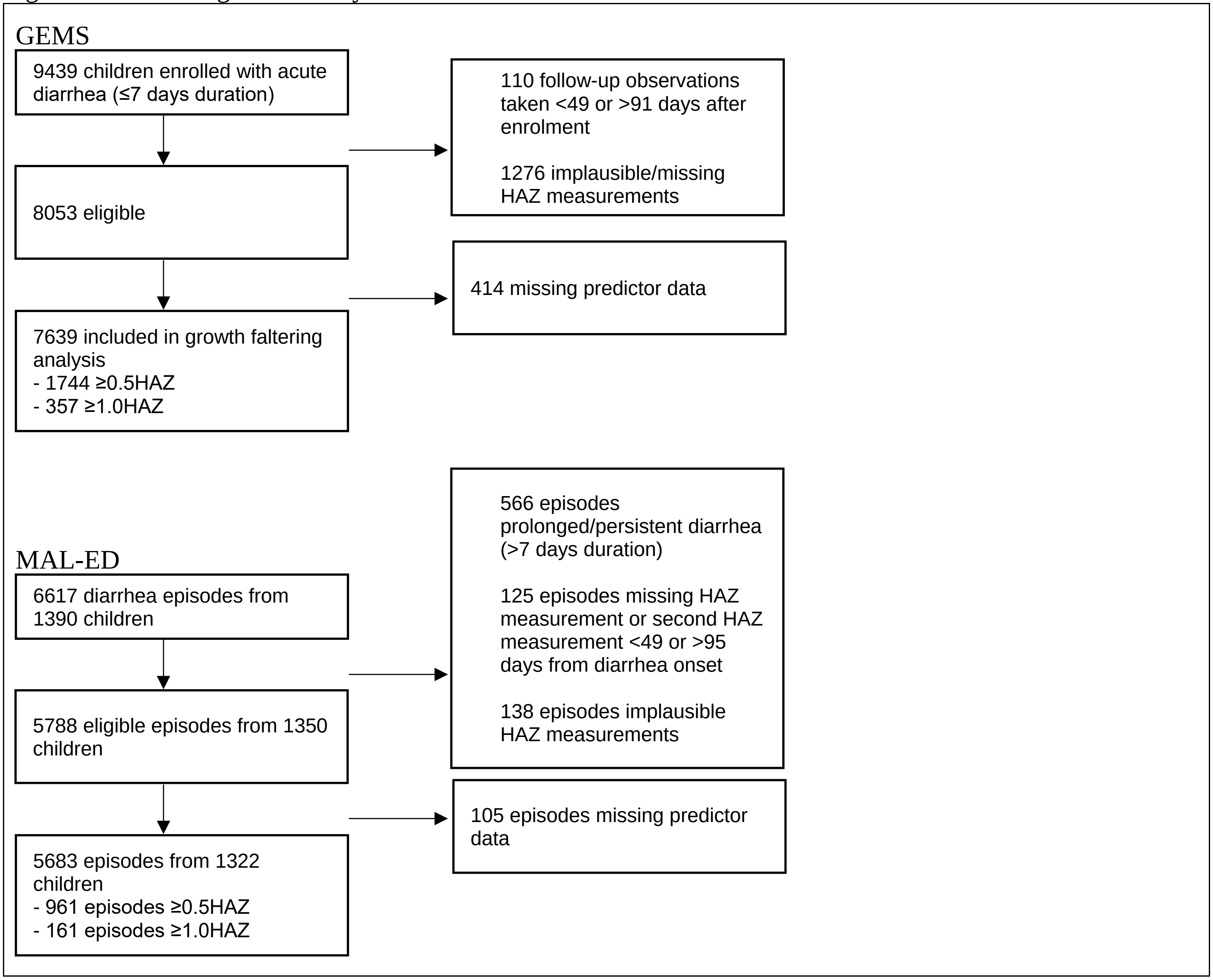

Table S1: Full list of considered predictor variables

| GEMS cases | GEMS controls | MAL-ED |
|---|---|---|
| Study site (site)<br>Child sex (f3_gender)<br>Loss of skin turgor (f3_drh_turgor)<br>Intravenous rehydration (f3_drh_iv)<br>Hospitalized (f3_drh_hosp)<br>Your relationship to the child (f4a_relationship)<br>Where child’s father lives (f4a_dad_live)<br>Primary caregiver’s max school (f4a_prim_schl)<br>People living in house 6 months (f4a_ppl_house)<br>Children under 60 months in the house (f4a_yng_children)<br>How many rooms used for sleeping (f4a_slp_rooms)<br>Predominant floor (f4a_floor)<br>Electricity (f4a_house_elec)<br>Bicycle/rickshaw (f4a_house_bike)<br>Telephone (f4a_house_phone)<br>Television (f4a_house_tele)<br>Car/truck (f4a_house_car)<br>Animal-drawn cart (f4a_house_cart)<br>Motorcycle/scooter (f4a_house_scoot)<br>Refrigerator (f4a_house_fridge)<br>Agricultural land (f4a_house_agland)<br>Radio (f4a_house_radio)<br>Boat with motor (f4a_house_boat)<br>None of the above assets (f4a_house_none)<br>Electricity (f4a_fuel_elec)<br>Biogas (f4a_fuel_biogas)<br>Straw/shrubs/grass (f4a_fuel_grass)<br>Liquid propane gas (f4a_fuel_propane)<br>Coal/lignite (f4a_fuel_coal)<br>Animal dung (f4a_fuel_dung)<br>Natural gas (f4a_fuel_natgas)<br>Charcoal (f4a_fuel_charcoal)<br>Agricultural crop residue (f4a_fuel_crop) | Study site (site)<br>Your relationship to the child (f7_relation)<br>Where child’s father lives (f7_dad_live)<br>Primary caregiver’s max school (f7_prim_schl)<br>People living in house 6 months (f7_ppl_house)<br>Children under 60 months in the house (f7_yng_childrn)<br>How many rooms used for sleeping (f7_slp_rooms)<br>Predominant floor (f7_floor)<br>Electricity (f7_house_elec)<br>Bicycle/rickshaw (f7_house_bike)<br>Telephone (f7_house_phone)<br>Television (f7_house_tele)<br>Car/track (f7_house_car)<br>Animal-drawn cart (f7_house_cart)<br>Motorcycle/scooter (f7_house_scoot)<br>Refrigerator (f7_house_fridge)<br>Agricultural land (f7_house_agland)<br>Radio (f7_house_radio)<br>Moat with motor (f7_house_boat)<br>None of the above assets (f7_house_none)<br>Electricity (f7_fuel_elec)<br>Biogas (f7_fuel_biogas)<br>Straw/shrubs/grass (f7_fuel_grass)<br>Liquid propane gas (f7_fuel_propane)<br>Coal/lignite (f7_fuel_coal)<br>Animal dung (f7_fuel_dung)<br>Natural gas (f7_fuel_natgas)<br>Charcoal (f7_fuel_charcoal)<br>Agricultural crop residue (f7_fuel_crop)<br>Kerosene (f7_fuel_kero)<br>Wood (f7_fuel_wood)<br>Other fuel (f7_fuel_other)<br>Goat (f7_ani_goat)<br>Sheep (f7_ani_sheep)<br>Dog (f7_ani_dog)<br>Cat (f7_ani_cat) | Cumulative diarrheal episode count (diar_epi_ct)<br>HAZ measurement no more than 31 days before onset of diarrhea (HAZ_1)<br>Diarrheal episode duration (days) (diar_dur)<br>Cumulative days within diarrheal episodes (diar_days_sum)<br>Dairrhea duration categorization (diar_dur_cat)<br>Max loose stools at episode (loose_stool_max)<br>Blood in stool at episode (blood)<br>Days with vomiting at episode (vomit_dur)<br>Days with decreased appetite at episode (app_dec_dur)<br>Max dehydration categorization (dehyd_max_cat)<br>Fever at episode categorization (fev_bin)<br>Cumulative days in this ALRI episode (ALRI_dur)<br>Age (days) (age)<br>Total days of breastfeeding (breast_totdays**)<br>Days since last diarrheal episode (diar_days_since)<br>ORT administered, caregiver report (ORT_caregiver)<br>Hospitalized (hosp)<br>Indrawing, fieldworker assessment (indraw_any)<br>Caregiver reported sleepiness (sleepy_any)<br>Caregiver reported difficult to awaken (unawake_any)<br>Caregiver reported use of antibiotics (abx_any)<br>Caregiver reported use of ORT (ORT_any)<br>If hospitalized that diarrhea episode (hosp_any)<br>First day of the diarrhea episode (first_diar_day)<br>Breastfed within first 24hr of birth (breast_24) |

| | | |
|---|---|---|
| Kerosene (f4a_fuel_kero)<br>Wood (f4a_fuel_wood)<br>Other fuel (f4a_fuel_other)<br>Goat (f4a_ani_goat)<br>Sheep (f4a_ani_sheep)<br>Dog (f4a_ani_dog)<br>Cat (f4a_ani_cat)<br>Cow (f4a_ani_cow)<br>Rodents (f4a_ani_rodents)<br>Fowl (f4a_ani_fowl)<br>Other animal (f4a_ani_other)<br>No animals (f4a_ani_no)<br>Water piped to house (f4a_water_house)<br>Covered well in house/yard (f4a_water_covwell)<br>Water piped into yard (f4a_water_yard)<br>Covered public well (f4a_water_covpwell)<br>Public tap (f4a_water_pubtap)<br>Protected spring (f4a_water_prospring)<br>Open well in house/yard (f4a_water_well)<br>Unprotected spring (f4a_water_unspring)<br>Open public well (f4a_water_pubwell)<br>River/stream (f4a_water_river)<br>Pond/lake (f4a_water_pond)<br>Deep tube well (f4a_water_deepwell)<br>Rainwater (f4a_water_rain)<br>Shallow tube well (f4a_water_shallwell)<br>Bought water (f4a_water_bought)<br>Other water source (f4a_water_othr)<br>Bore hole (f4a_water_bore)<br>Main source of drinking water (f4a_ms_water*)<br>How often is water available (f4a_water_avail)<br>Did you give the child stored water (f4a_store_water)<br>Do you usually treat drinking water? (f4a_trt_water)<br>Usual treatment method (f4a_trt_method) | Cow (f7_ani_cow)<br>Rodents (f7_ani_rodents)<br>Fowl (f7_ani_fowl)<br>Other animals (f7_ani_other)<br>No animals f7_ani_no<br>Water piped to house (f7_water_house)<br>Covered well in house/yard (f7_water_covwell)<br>Water piped into yard (f7_water_yard)<br>Covered public well (f7_water_covpwell)<br>Public tap (f7_water_pubtap)<br>Protected spring (f7_water_prospring)<br>Open well in house/yard (f7_water_well)<br>Unprotected spring (f7_water_unspring)<br>Open public well (f7_water_pubwell)<br>River/stream (f7_water_river)<br>Pond/lake (f7_water_pond)<br>Deep tube well (f7_water_deepwell)<br>Rainwater (f7_water_rain)<br>Shallow tube well (f7_water_shallwell)<br>Bought water (f7_water_bought)<br>Other water source (f7_water_othr)<br>Bore hole (f7_water_bore)<br>Main source of drinking water (f7_ms_water*)<br>How often is water available (f7_water_avail)<br>Did you give the child stored water? (f7_store_water)<br>Do you usually treat drinking water? (f7_trt_water) Do you usually treat drinking water? (f7_trt_method)<br>How are child's feces disposed (f7_disp_feces)<br>Facility use to dispose of feces (f7_fac_waste)<br>Wash hands before eating? (f7_wash_eat) | Time between birth and first breastfeeding (time_to_breast)<br>Fed colostrum (colostrum)<br>Prelacteal feeding (prelacteal)<br>Sex (sex)<br>Total days in all diarrheal episodes (tot_diar)<br>Drinking water source (water_source)<br>Persons sleeping in dwelling (ppl_slp)<br>Mean people per room (mean_ppl)<br>Improved/unimproved sanitation (sani_score)<br>Improve/unimproved drinking water (water_score)<br>Household has a bed (bed)<br>Household has a television (tv)<br>Household has a refrigerator (fridge)<br>Household has a table (table)<br>Household has a chai (chair)<br>Main material of roof (roof)<br>Main material of floor (floor)<br>Main material of walls (wall)<br>Household has a bank account (bank)<br>Kitchen located in a separate room (kitchen)<br>Years of formal education mother received (edu)<br>Mother ever attended formal schooling (edu2)<br>Total number of rooms in house (rm_ct)<br>Noniles of average monthly household income in USD (income_score)<br>Fewer than 2 people per room (two_ppl_rm)<br>Toilet/latrine has concrete floor (sani_concrete)<br>Toilet/latrine type (sani_type)<br>Country (country) |

| | |
|---|---|
| How are child's feces disposed (f4a_disp_feces)<br>Facility used to dispose of feces (f4a_fac_waste)<br>How many households share facility? (f4a_share_fac)<br>Wash hands before eating? (f4a_wash_eat)<br>Wash hands before cooking (f4a_wash_cook)<br>Wash hands before you nurse? (f4a_wash_nurse)<br>Wash hands after you defecate (f4a_wash_def)<br>Wash hands after handling animals (f4a_wash_animal)<br>Wash hands after cleaning a child (f4a_wash_child)<br>Wash hands other times (f4a_wash_othr)<br>What do you use to wash your hands? (f4a_wash_use)<br>Is the child currently breastfed? (f4a_breastfed)<br>How long as this diarrhea episode lasted (days)? (f4a_drh_days)<br>Maximum number of loose stools (f4a_max_stools)<br>Blood in stools (f4a_drh_blood)<br>Vomiting 3 or more times per day (f4a_drh_vomit)<br>Very thirsty (f4a_drh_thirst)<br>Drank much less than usual (f4a_drh_lessdrink)<br>Belly pain (f4a_drh_bellypain)<br>Irritable or restless (f4a_drh_restless)<br>Decreased activity or lethargy (f4a_drh_lethrgy)<br>Loss of consciousness (f4a_drh_consc)<br>Rectal straining (f4a_drh_strain)<br>Rectal prolapse (f4a_drh_prolapse)<br>Cough (f4a_drh_cough)<br>Convulsions (f4a_drh_conv)<br>Very thirsty (f4a_cur_thirsty)<br>Wrinkled skin (f4a_cur_skin)<br>Irritable or restless (f4a_cur_restless)<br>Dry mouth (f4a_cur_drymouth) | Wash hands before cooking? (f7_wash_cook)<br>Wash hands before you nurse (f7_wash_nurse)<br>Wash hands after you defecate? (f7_wash_def)<br>Wash hands after handling animals? (f7_wash_animal)<br>Wash hands after cleaning child? (f7_wash_child)<br>Wash hands other times? (f7_wash_othr)<br>What do you use to wash your hands? (f7_wash_use)<br>Is the child currently breastfeeding? (f7_breastfed)<br>Seek outside care? (f7_seekcare)<br>Length/height-for-age z-score (f7_haz)<br>Axillary temperature (f7_temp)<br>Calculated respiratory rate (f7_resp)<br>Bipedal edema (f7_bipedal)<br>Abnormal hair (f7_abn_hair)<br>Undernutrition (f7_under_nutr)<br>Skin as 'flaky paint' appearance (f7_skin_flaky)<br>Child age (months) (base_age) |

| |
|---|
| Fast breathing (f4a_cur_fastbreath)<br>ORALITE or ORS (f4a_hometrt_ors)<br>Homemade fluid (f4a_hometrt_maize)<br>Special mile or infant formula (f4a_hometrt_milk)<br>Home remedy/herbal medication (f4a_hometrt_herb)<br>Zinc (f4a_hometrt_zinc)<br>No special remedies given (f4a_hometrt_none)<br>Any other liquids (f4a_hometrt_othrliq)<br>Antibiotics (f4a_hometrt_ab)<br>Other treatment (f4a_hometrt_othr1)<br>Other treatment (f4a_hometrt_othr2)<br>How much offered to drink (f4a_offr_drink)<br>Seek outside care (f4a_seek_outside)<br>Pharmacy (f4a_seek_pharm)<br>Friend/relative (f4a_seek_friend)<br>Traditional healer (f4a_seek_healer)<br>Unlicensed practitioner (f4a_seek_doc)<br>Licensed practitioner (f4a_seek_privdoc)<br>Bought a remedy (f4a_seek_remdy)<br>Other hospital/center (f4a_seek_other)<br>Length/height-for-age z-score (f4b_haz)<br>Axillary temperature (f4b_temp)<br>Respiratory rate per minute (f4b_resp)<br>Chest indrawing (f4b_chest_indrw)<br>Eyes (f4b_eyes)<br>Mouth (f4b_mouth)<br>Skin pinch (f4b_skin)<br>Mental status (f4b_mental)<br>Rectal prolapse (f4b_rectal)<br>Bipedal edema (f4b_bipedal)<br>Abnormal hair (f4b_abn_hair)<br>Undernutrition (f4b_under_nutr) |

| |
|---|
| Skin as ‘flaky paint’ appearance (f4b_skin_flaky)<br>Staff observed a stool sample (f4b_observe_stool)<br>Nature of the stool (f4b_nature_stool)<br>Receive rehydration here (f4b_recommend)<br>Child was admitted to hospital (f4b_admit)<br>Child age (months) (base_age) |

*f4a_ms_water and f7_ms_water were recategorized into the following: surface, other unimproved, other improved, piped, other(1, 2)
**breast_totdays is calculated from the cumulative number of days of each of the following types of breastfeeding, combined using the listed formula: exclusive, predominant, partial, and none; breast_totdays=(breast_excl*1)+(breast_predom*0.75)+(breast_part*0.5)+(breast_not*0))

Table S2: Total sample size and growth faltering in GEMS by site

| N’s | The Gambia | Mali | Mozambique | Kenya | India | Bangladesh | Pakistan |
|---|---|---|---|---|---|---|---|
| ≥0.5 decrease in HAZ | 251 (31.9%) | 255 (14.9%) | 145 (34.5%) | 296 (28.4%) | 269 (18.5%) | 304 (23.1%) | 224 (24.9%) |
| ≥1.0 decrease in HAZ | 48 (6.1%) | 51 (3.0%) | 42 (10.0%) | 76 (7.3%) | 55 (3.8%) | 38 (2.9%) | 47 (5.2%) |
| Total | 788 | 1715 | 420 | 1042 | 1457 | 1316 | 901 |

Table S3: Total Sample size and growth faltering in GEMS by age

| N’s | 0-11mo | 12-23mo | 24-59mo |
|---|---|---|---|
| ≥0.5 decrease in HAZ | 1077 (33.7%) | 584 (22.3%) | 83 (4.6%) |
| ≥1.0 decrease in HAZ | 293 (9.2%) | 64 (2.4%) | 0 (0%) |
| Total | 3196 | 2622 | 1821 |

Figure S2: Number of variables and AUC for random forest regression and logistic regression

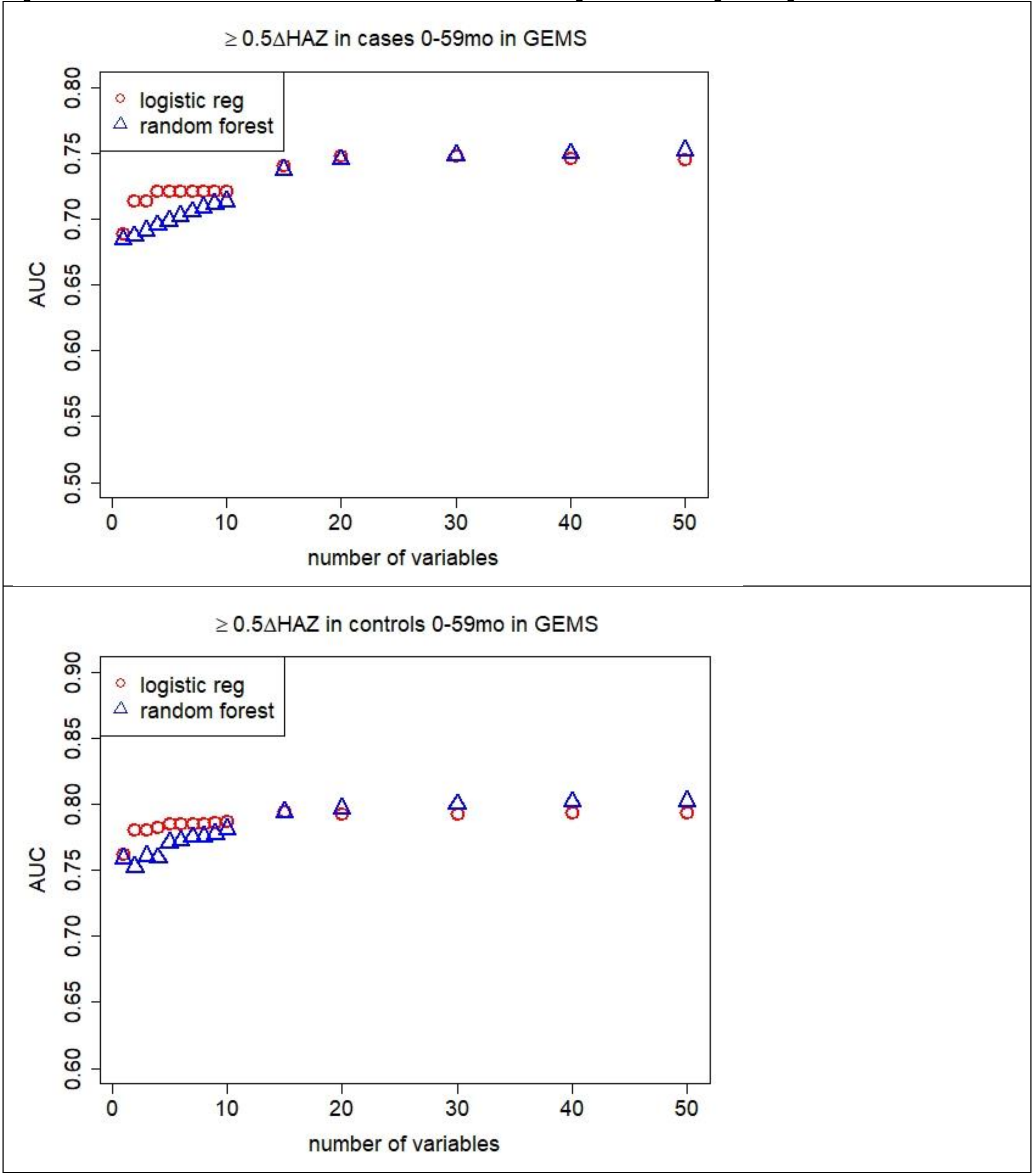

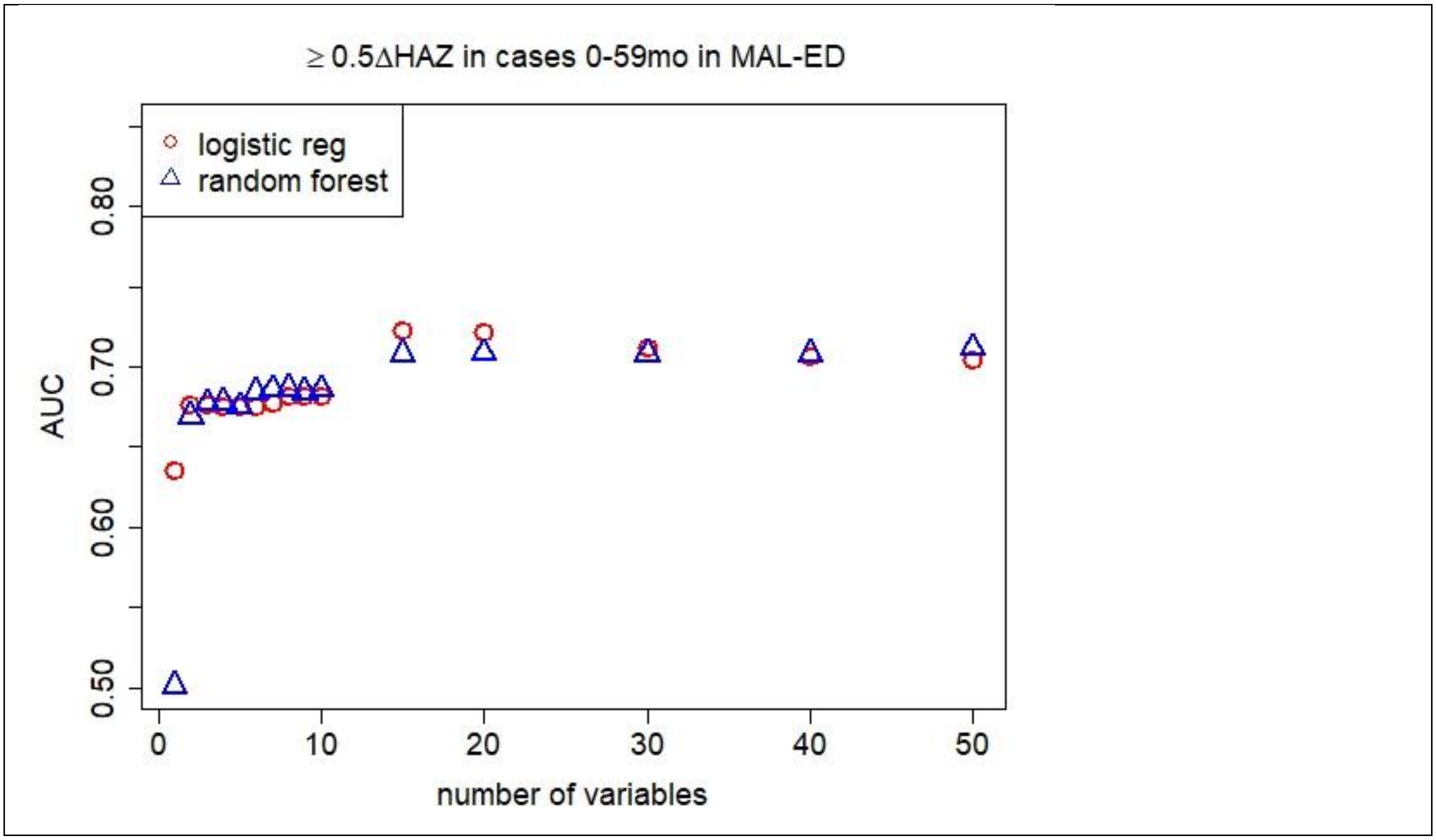

≥ 0.5ΔHAZ in cases 0-59mo in MAL-ED
logistic reg
random forest
AUC
0.50
0.60
0.70
0.80
0
10
20
30
40
50
number of variables

Table S4: Variable importance ordering, cross-validated average AUC, odds ratios, and 95% confidence intervals for logistic regression models predicting growth faltering (≥0.5 decrease in HAZ) in children 0-59mo in LMICs in GEMS and MAL-ED ranked from most to less predictive (highest to lower variance reduction)

| GEMS | | MAL-ED | |
|---|---|---|---|
| AUC (95% CI): 0.72 (0.719, 0.724) | | AUC (95% CI): 0.68 (0.67, 0.69) | |
| Variables | OR (95% CI) | Variables | OR (95% CI) |
| Age (months) | 0.92 (0.91, 0.93) | HAZ | 1.35 (1.17, 1.55) |
| HAZ | 1.30 (1.24, 1.36) | Age (days) | 1.00 (1.00, 1.00) |
| Respiratory rate | 1.00 (0.99, 1.00) | Total days breastfed | 1.00 (1.00, 1.01) |
| Temperature | 1.28 (1.21, 1.36) | Tot. days with diarrhea during study (tot_diar) | 0.98 (0.97, 1.00) |
| Num. ppl living in household | 1.00 (0.99, 1.01) | Mean num. people per room | 1.05 (0.94, 1.16) |
| Num. ppl sleeping in household | 1.00 (0.98, 1.03) | Num. days of diarrhea in lifetime at presentation (diar_days_sum) | 1.02 (0.99, 1.04) |
| Num. days of diarrhea at presentation | 0.98 (0.93, 1.02) | Maternal education (years) | 0.95 (0.91, 1.00) |
| Num. other households share fecal waste disposal facility | 0.99 (0.97, 1.00) | Num. days since last diarrhea episode (diar_days_since) | 1.00 (1.00, 1.00) |
| Currently breastfeeding | 0.85 (0.74, 0.97) | Num. ppl. sleep in dwelling | 1.07 (1.01, 1.12) |
| Num. children <60months live in household | 0.99 (0.95, 1.03) | Max. num. loose stools per day in this diarrhea episode | 0.93 (0.86, 1.00) |

Table S5: **GROWTH FALTERING:** Variable importance ordering and cross-validated average overall AUC and AUC by patient subset and 95% confidence intervals for a 5 (bold) and 10 (italicized) variable logistic regression model for predicting growth faltering in children derived in GEMS and MAL-ED data (≥0.5 decrease in HAZ)

| Data | GEMS | GEMS | MAL-ED | GEMS | MAL-ED | GEMS | MAL-ED | GEMS |
|---|---|---|---|---|---|---|---|---|
| Patient Subset | 0-59mo (main text model) | 0-11mo | 0-11mo | 12-23mo | 12-23mo | 0-23mo (for external validation) | 0-23mo | 24-59mo |
| AUCs | **0.72 (0.72, 0.72)** | **0.60 (0.59, 0.60)** | **0.60 (0.60, 0.61)** | **0.66 (0.65, 0.66)** | **0.65 (0.64, 0.66)** | **0.64 (0.63, 0.64)** | **0.62 (0.61, 0.63)** | **0.78 (0.77, 0.79)** |
| | *0.72 (0.71, 0.73)* | *0.60 (0.59, 0.60)* | *0.63 (0.62, 0.63)* | *0.70 (0.69, 0.70)* | *0.64 (0.63, 0.65)* | *0.64 (0.64, 0.64)* | *0.63 (0.62, 0.64)* | *0.76 (0.75, 0.77)* |
| 1 | Age (months) | HAZ | HAZ | HAZ | Age (days) | HAZ | HAZ | Temp |
| 2 | HAZ | Temp | Total days breastfeeding | Respiratory rate | HAZ | Age (months) | Age (days) | Respiratory rate |
| 3 | Respirator rate | Respiratory rate | Age (days) | Temp | Total days breastfeeding | Temperature | Total days breastfeeding | Age (months) |
| 4 | Temperature | Num. people living in household | Tot. days with diarrhea during study | Age (months) | Tot. days with diarrhea during study | Respiratory rate | Tot. days with diarrhea during study | HAZ |
| 5 | Num. people living in household | Age (months) | Mean num. of people per room | Num. people living in household | Num. days since last diarrhea episode | Num. people living in household | Mean num. people per room | Num. people living in household |
| 6 | Num. rooms used for sleeping | Num. other households share latrine | Num. days of diarrhea in lifetime at presentation | Num. rooms used for sleeping | Num. days of diarrhea in lifetime at presentation | Num. rooms used for sleeping | Num. days of diarrhea in lifetime at presentation | Num. other households that share same fecal waste facility |
| 7 | Num. days of diarrhea at presentation | Num. days of diarrhea at presentation | Num. ppl. sleep in dwelling | Received recommended rehydration at health center | Mean num. people per room | Num. days of diarrhea at presentation | Num. ppl. Sleep in dwelling | Num. days of diarrhea at presentation |
| 8 | Num. other households that share | Num. rooms used for sleeping | Max num. loose stools per day in this | Num. days of diarrhea at presentation | Maternal education (years) | Num. other households that share | Maternal education (years) | Num. rooms used for sleeping |

| | | | | | | | | |
|---|---|---|---|---|---|---|---|---|
| | same fecal waste facility | | diarrhea episode | | | same fecal waste facility | | |
| 9 | Breastfed | Num. children <60months live in household | Maternal education (years) | Num. other households that share same fecal waste facility | Num. ppl sleep in dwelling | Num. children <60 months live in household | Num. days since last diarrhea episode | Num. children <60months live in household |
| 10 | Num. children <60months live in household | Caregiver education | Num. days since last diarrhea episode | Num. children <60months live in household | Num. of diarrhea episodes in lifetime | Caregiver education | Avg. monthly household income in USD, noniles | Type of fecal waste facility used by household |

| Data | GEMS | GEMS | GEMS | GEMS | GEMS | GEMS |
|---|---|---|---|---|---|---|
| Patient Subset | 0-59mo (main text model) only HAZ considered | Only MUAC considered | HAZ + MUAC considered | main + month of diarrhea | Abx (b/f, during, rx, ever) | ≥1.0 ΔHAZ in 0-59mo |
| AUCs | **0.72 (0.72, 0.72)** | **0.70 (0.70, 0.70)** | **0.72 (0.72, 0.73)** | **0.72 (0.72, 0.72)** | **0.72 (0.72, 0.72)** | **0.80 (0.79, 0.80)** |
| | *0.72 (0.71, 0.73)* | *0.70 (0.70, 0.70)* | *0.72 (0.72, 0.73)* | *0.72 (0.72, 0.72)* | *0.72 (0.72, 0.72)* | *0.80 (0.79, 0.80)* |
| 1 | Age (months) | Age (months) | Age (months) | Age (months) | Age (months) | Age (months) |
| 2 | HAZ | MUAC | HAZ | HAZ | HAZ | HAZ |
| 3 | Respirator rate | Respiratory rate | MUAC | Respiratory rate | Respiratory rate | Respiratory rate |
| 4 | Temperature | Temperature | Respiratory rate | Temperature | Temperature | Temperature |
| 5 | Num. people living in household | Num. people living in household | Temperature | Num. people living in household | Num. people living in household | Num. people living in household |
| 6 | Num. rooms used for sleeping | Num. days of diarrhea at presentation | Num. people living in household | Month | Num. rooms used for sleeping | Num. other households that share same fecal waste facility |
| 7 | Num. days of diarrhea at presentation | Num. rooms used for sleeping | Num. rooms used for sleeping | Num. rooms used for sleeping | Num. days of diarrhea at presentation | Num. rooms used for sleeping |
| 8 | Num. other households that share same fecal waste facility | Num. other households that share same fecal waste facility | Num. days of diarrhea at presentation | Num. days of diarrhea at presentation | Num. other households that share same fecal waste facility | Num. days of diarrhea at presentation |

| | | | | | | |
|---|---|---|---|---|---|---|
| 9 | Breastfed | Breastfed | Num. other households that share same fecal waste facility | Num. other households that share same fecal waste facility | Breastfed | Num. children <60months live in household |
| 10 | Num. children <60months live in household | Num. children <60months live in household | Breastfed | Breastfed | Num. children <60months live in household | Breastfed |
| Rank of additional variable | N/A | N/A | N/A | 6th | 62nd (rx)<br>71st (during)<br>78th (ever)<br>84th (before) | N/A |

| Data Attributable fraction cutoff considered | GEMS | 0.3 | 0.3 | 0.3 | 0.3 | 0.5 | 0.5 | 0.5 | 0.5 | 0.7 | 0.7 | 0.7 | 0.7 |
|---|---|---|---|---|---|---|---|---|---|---|---|---|---|
| Patient Subset | 0-59mo (main text model) | main + Y/N Shigella* | main + Y/N crypto* | Main + Y/N Shigella + Y/N crypto* | main + Y/N any viral** | main + Y/N Shigella* | main + Y/N crypto* | Main + Y/N Shigella + Y/N crypto* | main + Y/N any viral** | main + Y/N Shigella* | main + Y/N crypto* | Main + Y/N Shigella + Y/N crypto* | main + Y/N any viral** |
| AUCs | **0.72 (0.72, 0.72)** | **0.73 (0.73, 0.74)** | **0.73 (0.73, 0.74)** | **0.73 (0.73, 0.74)** | **0.73 (0.73, 0.74)** | **0.73 (0.73, 0.74)** | **0.73 (0.73, 0.73)** | **0.73 (0.73, 0.74)** | **0.73 (0.73, 0.74)** | **0.73 (0.73, 0.74)** | **0.73 (0.73, 0.74)** | **0.73 (0.73, 0.74)** | **0.73 (0.73, 0.74)** |
| | *0.72 (0.71, 0.73)* | *0.73 (0.73, 0.74)* | *0.73 (0.73, 0.73)* | *0.73 (0.73, 0.74)* | *0.73 (0.73, 0.73)* | *0.73 (0.73, 0.73)* | *0.73, (0.73, 0.73)* | *0.73 (0.73, 0.74)* | *0.73 (0.73, 0.74)* | *0.73 (0.73, 0.73)* | *0.73, (0.73, 0.74)* | *0.73 (0.73, 0.73)* | *0.73 (0.73, 0.73)* |
| 1 | Age (months) | Age (months) | Age (months) | Age (months) | Age (months) | Age (months) | Age (months) | Age (months) | Age (months) | Age (months) | Age (months) | Age (months) | Age (months) |
| 2 | HAZ | HAZ | HAZ | HAZ | HAZ | HAZ | HAZ | HAZ | HAZ | HAZ | HAZ | HAZ | HAZ |
| 3 | Respirator rate | Resp rate | Resp rate | Resp rate | Resp rate | Resp rate | Resp rate | Resp rate | Resp rate | Resp rate | Resp rate | Resp rate | Resp rate |

| | | | | | | | | | | | | | |
|---|---|---|---|---|---|---|---|---|---|---|---|---|---|
| 4 | Temperature | Temperature | Temperature | Temperature | Temperature | Temperature | Temperature | Temperature | Temperature | Temperature | Temperature | Temperature | Temperature |
| 5 | Num. people living in household | Num. people living in household | Num. people living in household | Num. people living in household | Num. people living in household | Num. people living in household | Num. people living in household | Num. people living in household | Num. people living in household | Num. people living in household | Num. people living in household | Num. people living in household | Num. people living in household |
| 6 | Num. rooms used for sleeping | Num. days of diarrhea at presentation | Num. days of diarrhea at presentation | Num. days of diarrhea at presentation | Num. days of diarrhea at presentation | Num. days of diarrhea at presentation | Num. days of diarrhea at presentation | Breastfed | Num. days of diarrhea at presentation | Num. days of diarrhea at presentation | Num. days of diarrhea at presentation | Num. days of diarrhea at presentation | Breastfed |
| 7 | Num. days of diarrhea at presentation | Num. rooms used for sleeping | Breastfed | Num. rooms used for sleeping | Num. rooms used for sleeping | Breastfed | Breastfed | Num. days of diarrhea at presentation | Breastfed | Breastfed | Breastfed | Num. rooms used for sleeping | Num. days of diarrhea at presentation |
| 8 | Num. other households that share same fecal waste facility | Breastfed | Num. rooms used for sleeping | Breastfed | Breastfed | Num. rooms used for sleeping | Num. rooms used for sleeping | Num. rooms used for sleeping | Num. rooms used for sleeping | Num. rooms used for sleeping | Num. rooms used for sleeping | Breastfed | Num. rooms used for sleeping |
| 9 | Breastfed | Num. other households that share same fecal | Num. other households that share same fecal | Num. other households that share same fecal | Num. other households that share same fecal | Num. other households that share same fecal | Num. other households that share same fecal | Num. other households that share same fecal | Num. other households that share same fecal | Num. other households that share same fecal | Num. other households that share same fecal | Num. other households that share same fecal | Num. other households that share same fecal |

| | | | | | | | | | | | | | |
|---|---|---|---|---|---|---|---|---|---|---|---|---|---|
| | | waste facility | waste facility | waste facility | waste facility | waste facility | waste facility | waste facility | waste facility | waste facility | waste facility | waste facility | waste facility |
| 10 | Num. children <60months live in household | Num. children <60months live in household | Num. children <60months live in household | Num. children <60months live in household | Num. children <60months live in household | Num. children <60months live in household | Num. children <60months live in household | Num. children <60months live in household | Num. children <60months live in household | Num. children <60months live in household | Num. children <60months live in household | Num. children <60months live in household | Num. children <60months live in household |
| Rank of additional variable | n/a | 33rd | 18th | 19th (crypto) 30th (shigella) | 22nd | 49th | 20th | 22nd (crypto) 48th (shigella) | 26th | 60th | 56th | 52nd (crypto) 62nd (shigella) | 26th |

*n=4277 due to missing etiology data

**viral etiology include astrovirus, norovirus GII, rotavirus, sapovirus, and adenovirus, n=4277 due to missing etiology data

Figure S3: Calibration Curves

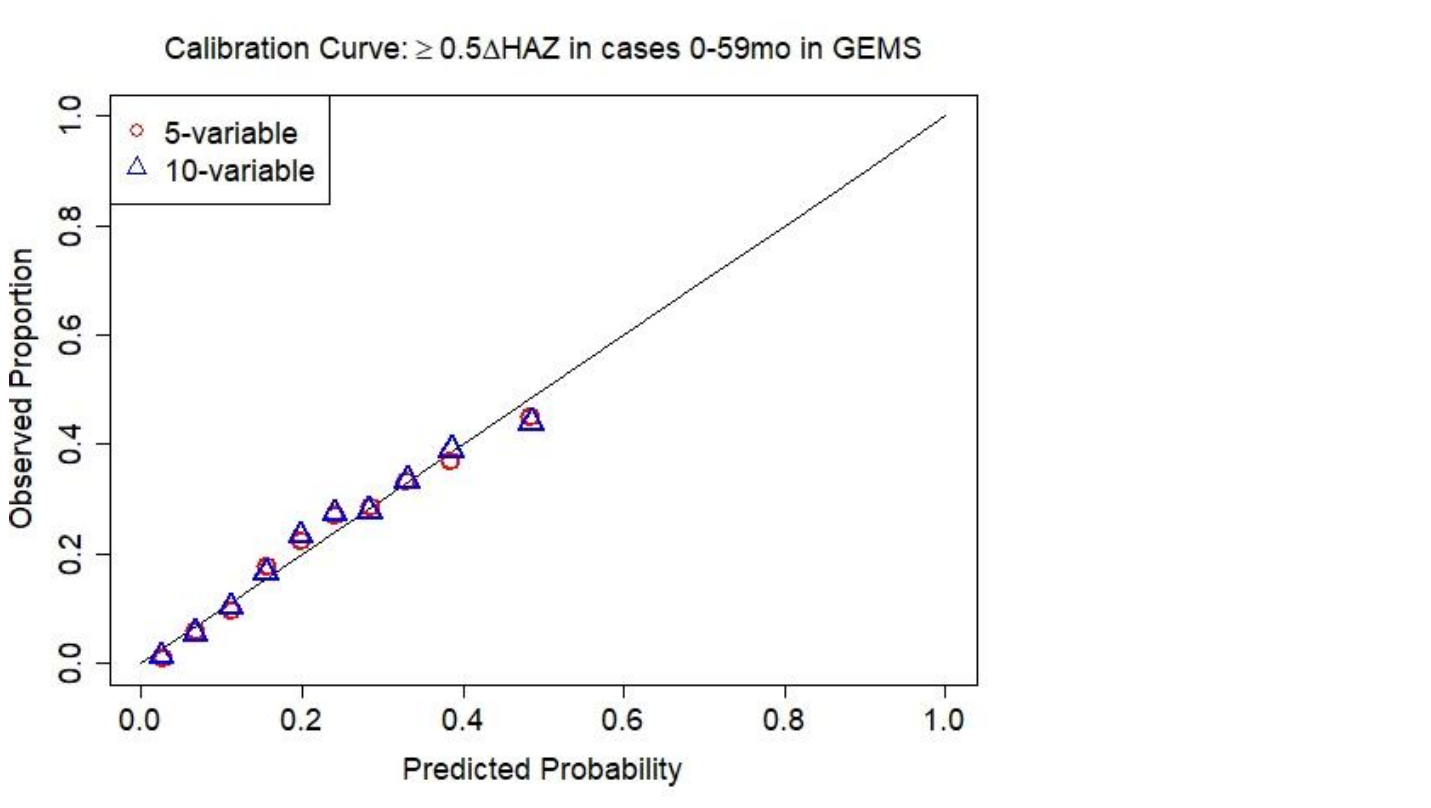

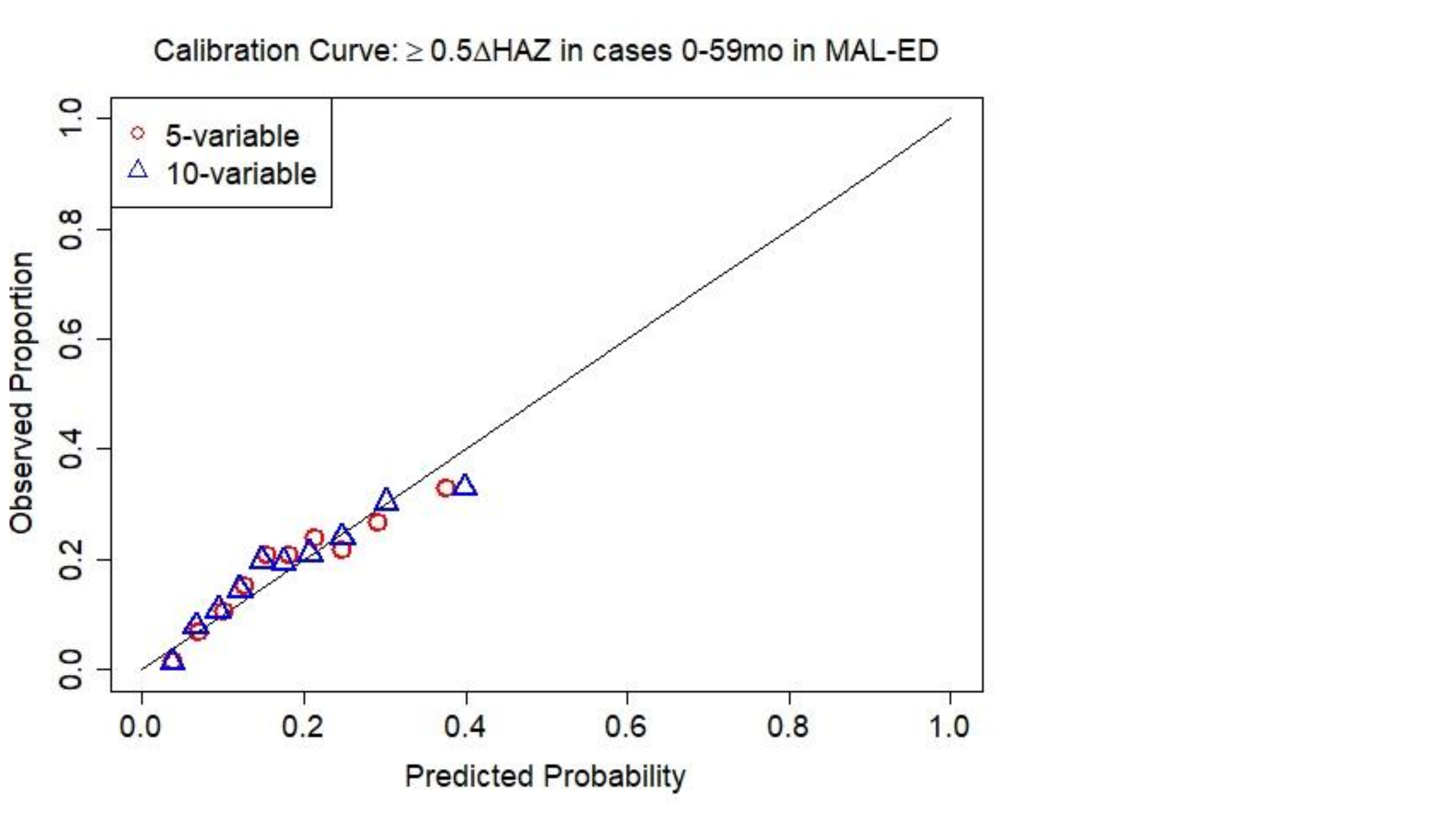

Figure S4

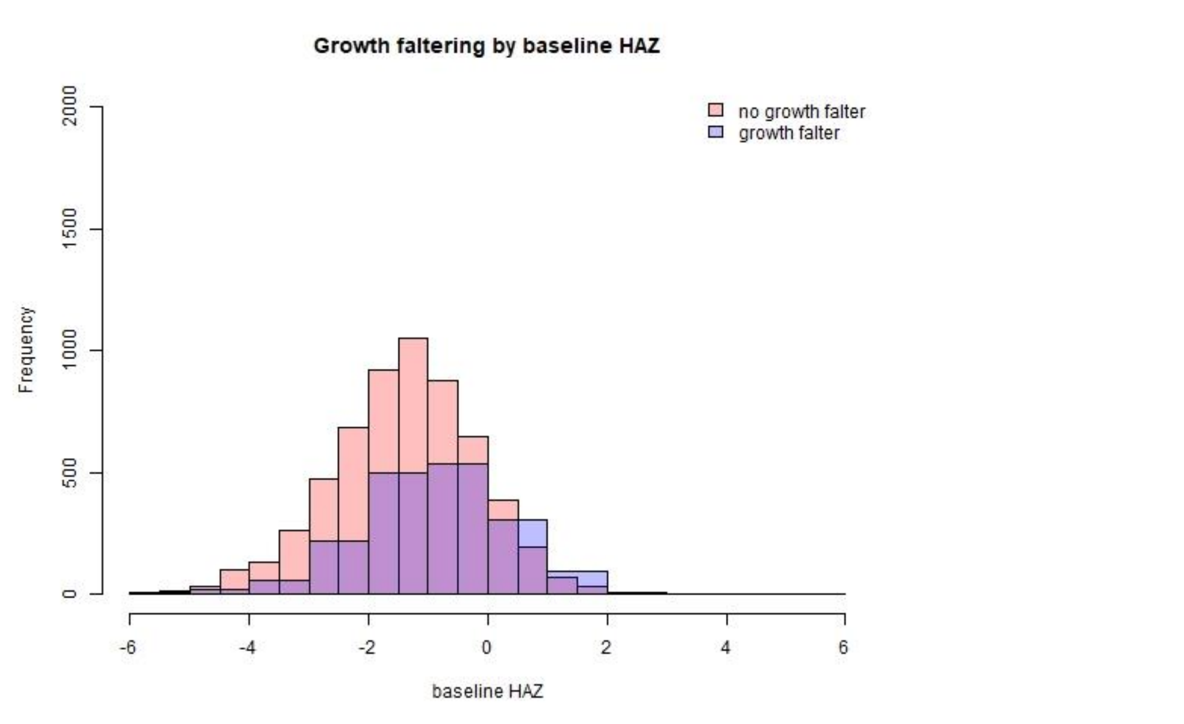

Mean baseline HAZ among children who did not growth falter: -1.34
Mean baseline HAZ among children who did growth falter: -0.84
Welch two sample t-test for difference in means $p<0.01$
